# Impact of SARS-CoV-2 infection on disease trajectory in youth with T1D: An EHR-based cohort study from the RECOVER program

**DOI:** 10.1101/2022.11.03.22281916

**Authors:** Priya Prahalad, Vitaly Lorman, Qiong Wu, Hanieh Razzaghi, Yong Chen, Nathan Pajor, Abigail Case, Seuli Bose-Brill, Jason Block, Payal B Patel, Suchitra Rao, Asuncion Mejias, Christopher B. Forrest, L. Charles Bailey, Ravi Jhaveri, Deepika Thacker, Dimitri A. Christakis, Grace M. Lee, the RECOVER Consortium

## Abstract

**Background:** Post-acute sequelae of SARS-Co-V-2 infection (PASC) is associated with worsening diabetes trajectory. It is unknown whether PASC in children with type 1 diabetes (T1D) manifests as worsening diabetes trajectory.

**Objective:** To explore the association between SARS-CoV-2 infection (COVID-19) and T1D-related healthcare utilization (for diabetic ketoacidosis [DKA] or severe hypoglycemia [SH]) or Hemoglobin (Hb) A1c trajectory.

**Methods:** We included children <21 years with T1D and ≥1 HbA1c prior to cohort entry, which was defined as COVID-19 (positive diagnostic test or diagnosis code for COVID-19, multisystem inflammatory syndrome in children, or PASC) or a randomly selected negative test for those who were negative throughout the study period (Broad Cohort). A subset with ≥1 HbA1c value from 28-275 days after cohort entry (Narrow Cohort) was included in the trajectory analysis. Propensity score-based matched cohort design followed by weighted Cox regression was used to evaluate the association of COVID-19 with healthcare utilization ≥28 days after cohort entry. Generalized estimating equation models were used to measure change in HbA1c in the Narrow cohort.

**Results:** From 03/01/2020-06/22/2022, 2,404 and 1,221 youth met entry criteria for the Broad and Narrow cohorts, respectively. The hazard ratio for utilization was (HR 1.45 [95%CI,0.97,2.16]). In the Narrow Cohort, the rate of change (slope) of HbA1c increased 91-180 days after cohort entry for those with COVID-19 (0.138 vs. -0.002, p=0.172). Beyond 180 days, greater declines in HbA1c were observed in the positive cohort (-0.104 vs. 0.008 per month, p=0.024).

**Conclusion:** While a trend towards worse outcomes following COVID-19 in T1D patients was observed, these findings were not statistically significant. Continued clinical monitoring of youth with T1D following COVID-19 is warranted.

**Authorship Statement:** Authorship has been determined according to ICMJE recommendations

**Disclaimer:** The content is solely the responsibility of the authors and does not necessarily represent the official views of the RECOVER Program, the NIH or other funders.

**Funding Source:** ⍰This research was funded by the National Institutes of Health (NIH) Agreement OT2HL161847-01 as part of the Researching COVID to Enhance Recovery (RECOVER) program of research.

## Introduction

Early in the COVID-19 pandemic, diabetes was prognostic of worse outcome (1; 2). Several reports from hospitalized patients have shown increased hyperglycemia in people with and without diabetes during the acute phase of infection with Severe Acute Respiratory Syndrome Coronavirus 2 (SARS-CoV-2) (3-5). The risk of developing long-term sequelae following infection with SARS-CoV-2 is also increased in adults with chronic medical conditions, including diabetes (6). Post-acute sequelae of SARS CoV-2 infection (PASC) is defined as new, continuing, or recurring symptoms that occur 4 or more weeks after initial coronavirus infection(7). In adults, symptoms of PASC can manifest as fatigue or symptoms affecting the cardiovascular, respiratory, neurologic, gastrointestinal, or musculoskeletal systems(8; 9). Reports suggest that 4.5% to 50% of adults with prior SARS-CoV-2 infection may develop symptoms of PASC(10-12). A similarly wide range of estimates have been noted for children (2% to 66%), though likely closer to ∼4% based on large population-based studies(13-16).

In children, a recent analysis of electronic health record (EHR) data suggests that PASC features can include symptoms such as changes in smell/taste, hair loss, and chest pain, and the most commonly associated conditions included myocarditis, respiratory distress and myositis(17). Prior data from the NIH RECOVER electronic health record (EHR) database shows that children with chronic medical conditions had higher rates of healthcare utilization 28 days or more after SARS-CoV-2 infection, compared to children without infection (18). However, the reasons for increased utilization were not well understood. Increased utilization may be due to symptoms associated with PASC (i.e., unrelated to their pre-existing conditions) or may be attributable to worsening of underlying chronic conditions (19).

Much of the previous data on PASC and diabetes has been focused on the incidence of new onset diabetes as a feature of PASC. Data from the Centers for Disease Control and US Department of Veterans Affairs show an increase in the incidence of new diabetes diagnoses in people with a history of COVID-19 infection(20; 21). However, these reports do not differentiate between type 1 and type 2 diabetes. Internationally, there have been mixed reports on increased incidence of diabetes, specifically type 1 diabetes (T1D in children, during the COVID-19 pandemic(22-25). A recent study of international EHR records showed an increase in the incidence of T1D in youth with a history of SARS-Co-V-2 infection(26) while an international registry study showed no change in the rare of rise of T1D(27). Since the SARS-CoV-2 virus is thought to infect beta cells(28; 29), new onset diabetes or worsening of disease trajectory in people with preexisting diabetes may both be features of PASC. In adults, there is data to suggest that SARS-CoV-2 infection can worsen dysglycemia in those with pre-existing diabetes(30).

In this manuscript, we report on the impact of SARS-CoV-2 infection on disease trajectory in children with preexisting type 1 diabetes (T1D). We selected T1D since it is associated with an increased risk of severe disease in children with COVID-19 infection (3-5; 31) and also thought to be a feature of PASC itself(20; 21). We focused on T1D since it is more common in the pediatric age group than type 2 diabetes (32-35). Using the NIH Researching COVID to Enhance Recovery (RECOVER) EHR database, we evaluated the impact of SARS-CoV-2 infection on T1D disease trajectory as measured by rates of hospital and emergency department utilization due to diabetes complications as well as changes in HbA1c trajectory, in youth with established type 1 diabetes.

## Materials and Methods

### Study Population

This study is part of the NIH RECOVER initiative which seeks to understand, treat, and prevent the post-acute sequalae of SARS-CoV-2 infection (PASC). Electronic health record (EHR) data was obtained from participating institutions in PEDSnet, a national network of pediatric healthcare systems that share clinical data to conduct observational research, clinical trials, and population surveillance. EHR data from inpatient, outpatient, emergency department, and administrative encounters within each participating health system were standardized to the PEDSnet common data model, an extension of the Observational Medical Outcomes Partnership common data model. Participating institutions for this study included Children’s Hospital of Philadelphia, Cincinnati Children’s Hospital Medical Center, Children’s Hospital of Colorado, Ann & Robert H. Lurie Children’s Hospital of Chicago, Nationwide Children’s Hospital, Nemours Children’s Health System (includes sites in Delaware and Florida), Seattle Children’s Hospital, and Stanford Children’s Health. Annually, these institutions provide service to 3.3% of the nation’s children. The RECOVER COVID-19 Database Version 2022-07-21 was used.

The Children’s Hospital of Philadelphia’s institutional review board designated this study as not human subjects research and the need for consent was waived.

### Study Design

In this retrospective, observational, matched cohort study, we included patients in the PEDSnet database who had a test, vaccine, or respiratory illness starting in January 1, 2020. Inclusion criteria were: <21 years of age with a diagnosis of T1D, presence of ≥1 HbA1C measurement in the 7 days to 3 years before cohort entry, and the presence of SARS-CoV-2 diagnostic testing or a diagnosis code for COVID-19, multi-system inflammatory syndrome in children (MIS-C), or PASC between March 1, 2020, and December 22, 2021. We defined a cohort of T1D patients with incident SARS-CoV-2 infection by presence of a positive SARS-CoV-2 PCR or antigen test performed in an outpatient, emergency department (ED), or inpatient setting, presence of a diagnosis term for multi-system inflammatory syndrome in children (MIS-C) or post-acute sequelae of COVID-19 at an outpatient, ED, or inpatient setting, or presence of any specific diagnosis term for COVID-19 associated with an inpatient or ED encounter. The cohort entry date was defined as the date of the earliest positive test or diagnosis. The comparison cohort was defined by presence of a negative SARS-CoV-2 viral test result, with no positive test result or diagnosis over the study period. In the comparison cohort, the cohort entry date was defined as the date of a randomly selected negative test for patients who had multiple negative tests (“Broad Cohort”). Patients with at least 6 months of follow-up time were included in our analyses.

To account for potential confounders, for both cohorts we used subclassification matching which grouped patients into 6 subclasses by propensity score and assigned weights to each subclass to balance age group, sex, race/ethnicity, HbA1c group (< 7%, 7-9%, ≥ 9%), insurance class, center, testing location, and cohort entry month in the two cohorts(36). After assigning patients the subclassification weights aweighted standardized mean differences (wSMDs) were computed to assess covariate balance.

### Calculation of Worsening Disease Trajectory

We examined worsening disease trajectory in two different ways. First, we developed a composite outcome to evaluate worsening disease trajectory in pediatric patients with T1D. In the Broad Cohort, the composite outcome was defined as an emergency department visit or hospitalization for diabetic ketoacidosis (DKA) or severe hypoglycemia (SH). Conditional hazard ratios were calculated using a Cox proportional hazards model (with weights from subclassification matching used to fit a weighted regression) to assess whether SARS-CoV-2 infection is associated with an increased hazard of hospitalization or emergency department visits for diabetic ketoacidosis or hypoglycemia. The adjusted hazard ratio (marginal estimate) quantifies the effect of infection of COVID-19 averaged in the weighted population. We also conducted a sensitivity analysis that did not require 6 months of follow-up time to explore whether time period of cohort entry (pre-Omicron compared to during the Omicron wave) modified the effect of COVID-positivity on our outcomes. Employing the same subclassification matching approach as above, we ran a proportional hazards model regressing on SARS-CoV-2 positivity, Omicron period, and an interaction term between SARS-Co-V-2 positivity and the Omicron period (defined as cohort entry on or after January 1, 2021).

Next, to further characterize whether disease trajectory following SARS-CoV-2 infection was altered in patients with T1D, we also evaluated the trajectory of HbA1c measurements using a “Narrow Cohort” of patients who had an additional HbA1c values from 28 days to 275 days after cohort entry. A longer time window was used to assess longer term effects on HbA1c outcomes. The propensity score model was fitted with the same variables described above, and 6 subclasses were created with covariate balance achieved. Within each propensity score stratum, a longitudinal regression model using the generalized estimating equation with autoregressive correlation structure was fitted to measure the change of HbA1c in positive and negative groups. The nonlinear change of HbA1c measurements was characterized by dividing the evaluation period into linear segments (i.e., prior to -90 days, -90 to -7 days, 28 to 90 days, 91 to 180 days, and later than 180 days). We further included SARS-CoV-2 positivity and its interactions with time segments after cohort entry as covariates. A fixed-effect meta-analysis was then used to aggregate effect sizes across propensity score strata (i.e., inverse-variance weighting). Unlike traditional longitudinal data, HbA1c measurements are not consistently observed for all subjects at set intervals, which results in an imbalanced data structure. In addition, the observation times may be associated with the underlying outcomes (HbA1c). For example, patients with HbA1c measurements available every 3 months may be more likely to have well-controlled diabetes compared to patients who do not have HbA1c data available. Hence, we conducted a sensitivity analysis to confirm the robustness of our findings. Specifically, the irregular visiting times were firstly modeled, and the generalized estimating equation (GEE) is then fitted with inverse-intensity-of-visit-process weighting (22; 23). The effect sizes in propensity score strata were then aggregated using fixed-effect meta-analysis. This sensitivity analysis is referred to as “Weighted GEE” (37; 38).

## Results

### Participant Characteristics

During the evaluation period (March 1, 2020, to June 22, 2022), there were a total of 2,404 youth in the Broad Cohort with at least 6 months of follow-up after cohort entry (Table 1a). Among patients in the Broad Cohort, 15.2% had a positive COVID-19 test at the time of cohort entry. A total of 1,221 individuals met criteria for inclusion in the Narrow Cohort (Table 1b). Similar to the Broad Cohort, 15.8% had a positive COVID-19 test at the time of cohort entry.

**Table 1.** Participant Characteristics in the (A) Broad Cohort and (B) Narrow cohort prior to matching (A)

|  | **Overall** | **Negative Cohort** | **Positive Cohort** |
| --- | --- | --- | --- |
| **n** | 2,404 | 2,039 | 365 |
| **Age at Cohort Entrance, n (%)** |  |  |  |
| < 5 years | 127 (5.2) | 115 (5.6) | 12 (3.3) |
| 5-11 years | 664 (27.6) | 591 (29.0) | 73 (20.0) |
| 12-15 years | 808 (33.6) | 669 (32.8) | 139 (38.1) |
| 16-20 years | 805 (33.5) | 664 (32.6) | 141 (38.6) |
| **Sex, n (%)** |  |  |  |
| Female | 1224 (50.9) | 1044 (51.2) | 180 (49.3) |
| Male/Other/Unknown | 1,180 (49.0) | 995 (48.7) | 186 (50.7) |
| **Race/Ethnicity, n(%)** |  |  |  |
| White Non-Hispanic | 1,484 (61.7) | 1,283 (62.9) | 201 (55.1) |
| Black Non-Hispanic | 361 (15.0) | 286 (14.0) | 75 (20.5) |
| Hispanic | 340 (14.1) | 281 (13.8) | 59 (16.2) |
| Other/Unknown | 219 (9.1) | 189 (9.2) | 30 (8.2) |
| **Insurance class, n(%)** |  |  |  |
| Public | 1,082 (45.0) | 901 (44.2) | 181 (49.6) |
| Private | 775 (32.2) | 685 (33.6) | 90 (24.7) |
| Other/Unknown | 547 (22.8) | 453 (22.2) | 94 (25.8) |
| **Cohort Entry Period\*** |  |  |  |
| Original strain (Mar 2020-Apr 2021) | 1,429 (59.4) | 1,208 (59.2) | 221 (60.5) |
| Alpha (May-Jul 2021) | 276 (11.5) | 258 (12.7) | 18 (4.9) |
| Delta (Aug-Dec 2021) | 699 (29.1) | 573 (28.1) | 126 (34.5) |
| **Test Location, n(%)** |  |  |  |
| Emergency Department | 667 (27.7) | 520 (25.5) | 147 (40.3) |
| Inpatient | 540 (22.5) | 486 (23.8) | 54 (14.8) |
| Outpatient Office | 436 (18.1) | 380 (18.6) | 56 (15.3) |
| Outpatient: Test Only | 761 (31.7) | 653 (32.0) | 108 (29.6) |

\*Our primary analyses required at least 6 months of follow-up after cohort entry.

**(B)**
|  | **Overall** | **Negative Cohort** | **Positive Cohort** |
| --- | --- | --- | --- |
| **n** | 1,221 | 1,028 | 193 |
| **Age at Cohort Entrance, n(%)** |  |  |  |
| < 5 years | 70 (5.8) | 64 (6.2) | 6 (3.1) |
| 5-11 years | 371 (30.4) | 331 (32.2) | 40 (20.7) |
| 12-15 years | 434 (35.5) | 347 (33.8) | 87 (45.1) |
| 16-20 years | 346 (28.3) | 286 (27.8) | 60 (31.1) |
| **Sex, n(%)** |  |  |  |
| Female | 605 (49.5) | 513 (49.9) | 92 (47.7) |
| Male | 616 (50.5) | 515 (50.1) | 101 (52.3) |
| **Race/Ethnicity, n(%)** |  |  |  |
| Non-Hispanic White | 774 (63.4) | 663 (64.5) | 111 (57.5) |
| Non-Hispanic Black | 184 (15.1) | 146 (14.2) | 38 (19.7) |
| Hispanic | 168 (13.8) | 138 (13.4) | 30 (15.5) |
| Other/Unknown | 95 (12.2) | 81 (7.9) | 14 (7.3) |
| **Insurance class, n(%)** |  |  |  |
| Public | 541 (44.3) | 449 (43.7) | 92 (47.7) |
| Private | 356 (29.2) | 315 (30.6) | 41 (21.2) |
| Other/unknown | 324 (26.5) | 264 (25.7) | 60 (31.1) |
| **Cohort Entry Period** |  |  |  |
| Original strain (Mar 2020-Apr 2021) | 708 (58.0) | 597 (58.1) | 111 (57.5) |
| Alpha (May-Jul 2021) | 140 (11.5) | 133 (12.9) | 7 (3.6) |
| Delta (Aug-Dec 2021) | 373 (30.5) | 298 (29.0) | 75 (38.9) |
| **Test Location, n(%)** |  |  |  |
| Emergency Department | 349 (28.6) | 277 (26.9) | 72 (37.3) |
| Inpatient | 238 (19.5) | 211 (20.5) | 27 (14.0) |
| Outpatient Office | 232 (19.0) | 197 (19.2) | 35 (18.1) |
| Outpatient: Test Only | 402 (32.9) | 343 (33.4) | 59 (30.6) |

### COVID-19 Infection and Outcomes

We used subclassification matching to balance age group, sex, race/ethnicity, HbA1c group (< 7%, 7-9%, ≥ 9%), insurance class, center, testing location, and cohort entry month in the two cohorts. The Broad cohort was well-balanced across all covariates after matching (|SMD|<0.1 for all covariates in the Broad cohort, except one month of cohort entry during which the |SMD| was <0.2, Figure 1).

**Figure 1.**
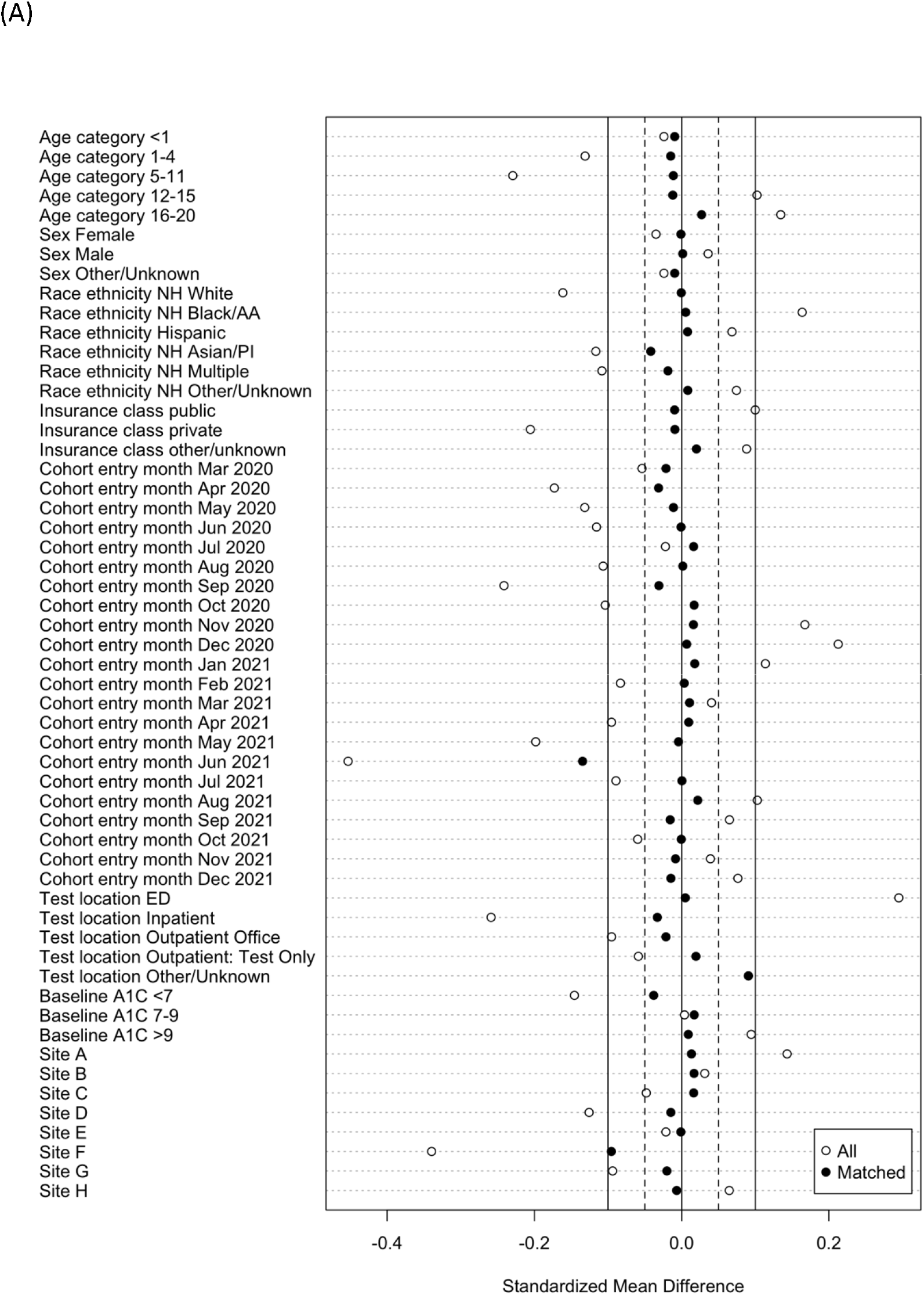
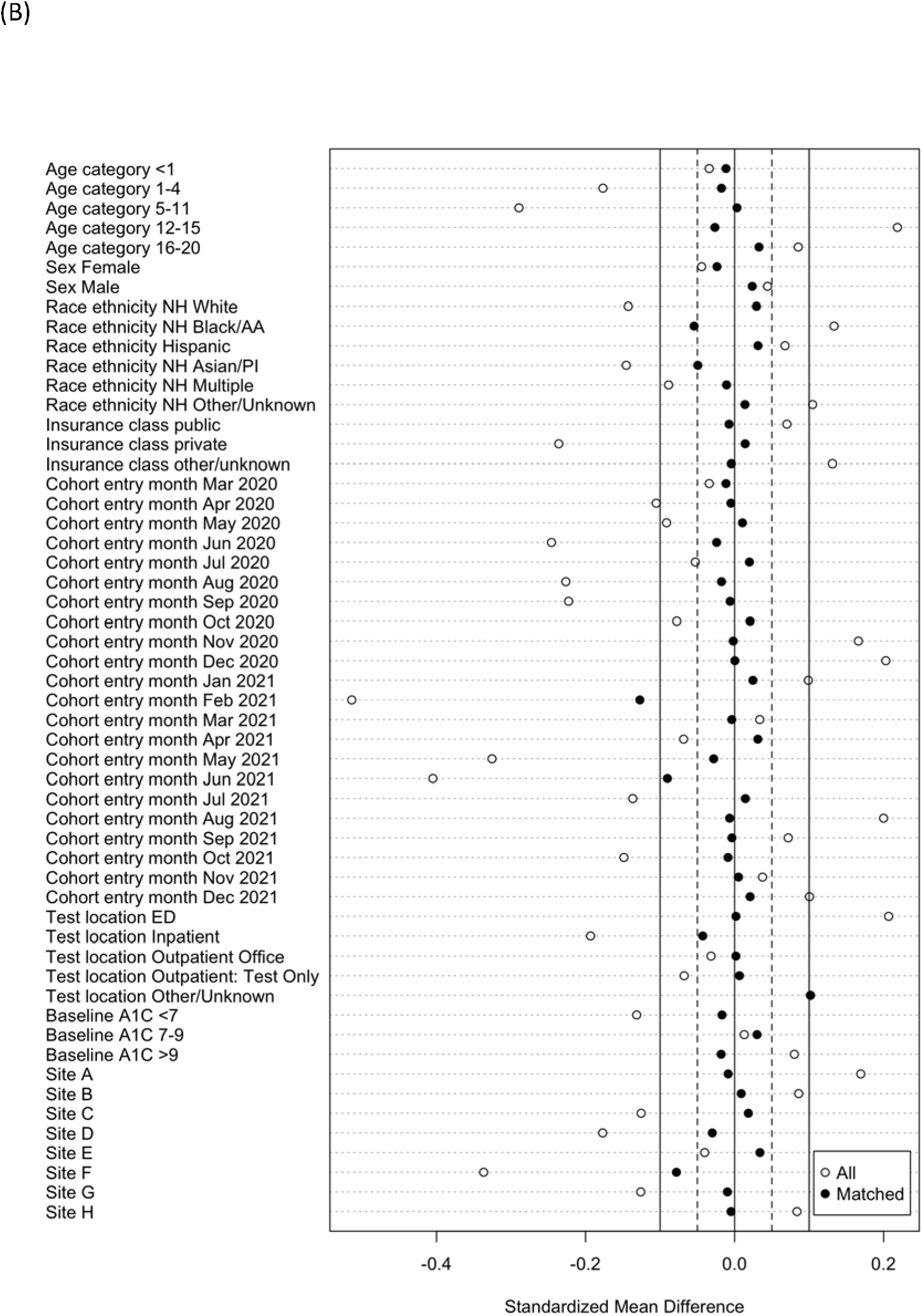
Propensity-score subclassification matching in the (A) broad and (B) narrow cohorts

The hazard ratio for our composite outcome in the Broad cohort, an emergency department visit or hospitalization for diabetic ketoacidosis or severe hypoglycemia ≥ 28 days after cohort entry, is 1.45 [95%CI: 0.97, 2.16]. This shows a trend towards increasing healthcare utilization for severe complications of T1D occurring ≥28 days after cohort entry, but it does not reach statistical significance. Kaplan Meier estimates of event-free survival are shown in Figure 2. In our sensitivity analysis, we did not require 6 months of follow-up time in order to capture information on patients with Omicron infection. The hazard ratio of our composite outcome occurring prior to the Omicron period (cohort entry 03/01/2020 to 12/31/2021) was 1.62 [95%CI:1.1,2.39] and during the Omicron period (01/01/2022) it decreased to 0.64 [95%CI (0.22, 1.85).

**Figure 2.**
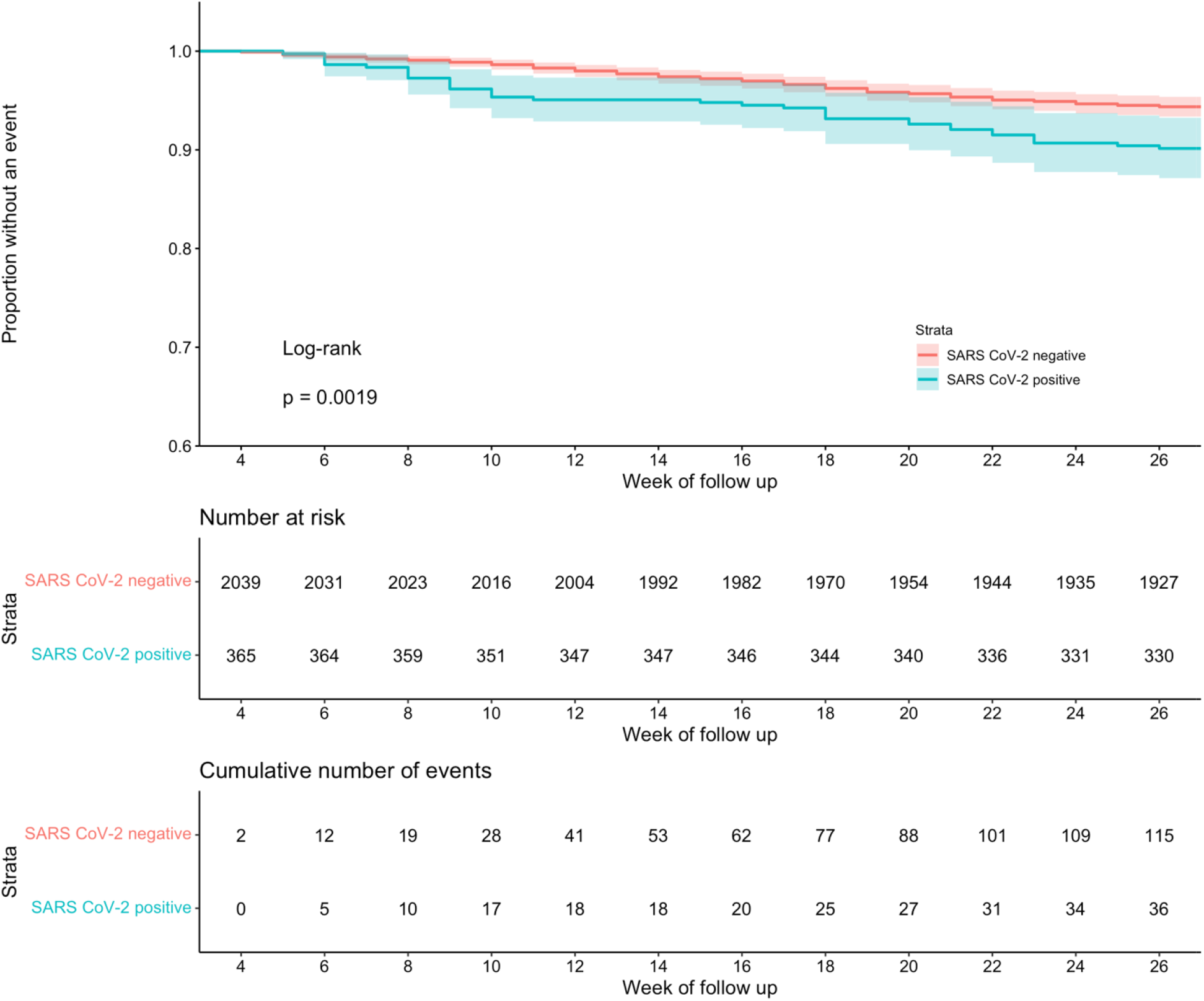
Kaplan Meier estimates of event-free survival for ED visit or hospitalization for diabetic ketoacidosis (DKA) or severe hypoglycemia (SH).

### HbA1c Trajectory Following COVID-19 Infection

Based on the aggregated regression coefficients, HbA1c trajectory was plotted starting from 6 months prior to cohort entry to 9 months after cohort entry for both positive and negative groups (Table 2, Figure 3). After propensity score stratification, the baseline HbA1c measures (at cohort entry) were similar for the two groups (i.e., the marginal effect of COVID-19 positivity is insignificant at the time of cohort entry). In those with a positive COVID-19 test or diagnosis, the rate of change (i.e., slope) of HbA1c values increased at day 90 after cohort entry (slope changed from -0.039 to 0.138 per month), however the increase of slope was not significant compared to the negative cohort (p=0.172). Beyond 180 days, HbA1c values had a greater rate of decline in the positive cohort (slope as -0.104 per month), and this change in trend at day 180 was statistically significant (p-value 0.024). The results from weighted GEE also showed a similar trajectory pattern with a statistically significant change in trend on day 180, which confirms the robustness of our findings.

**Table 2.**
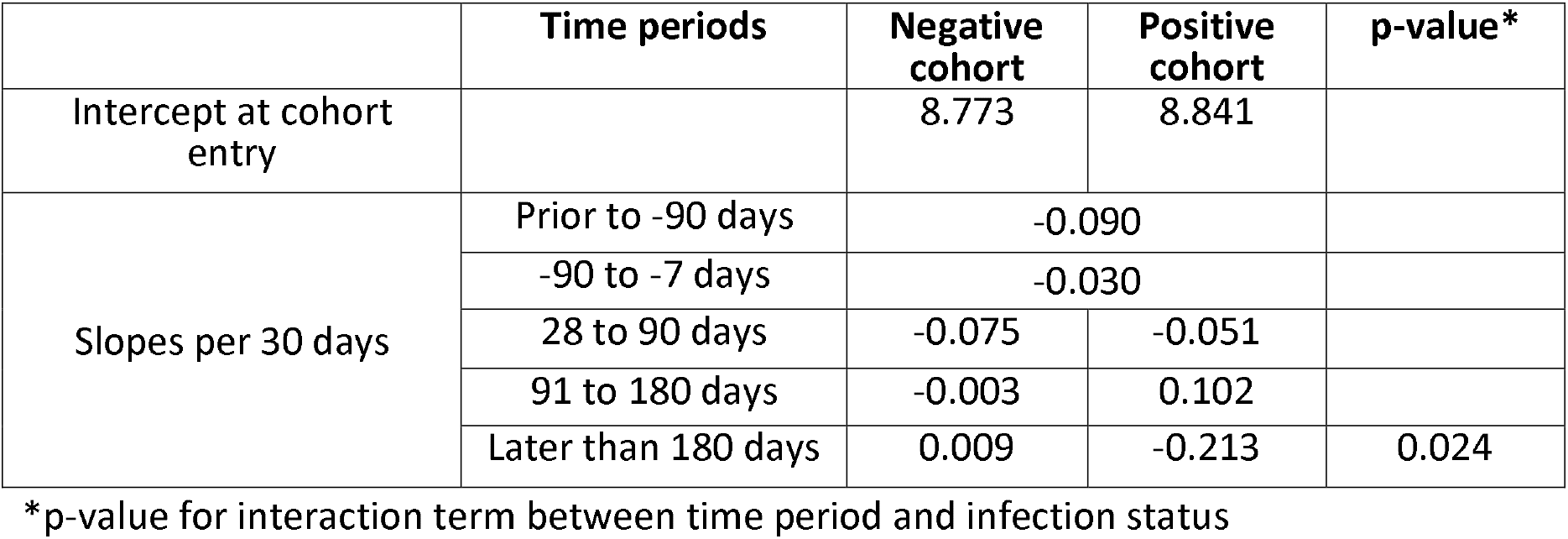
HbA1c trajectories measured as slopes during time periods before and after cohort entry.

**Figure 3.**
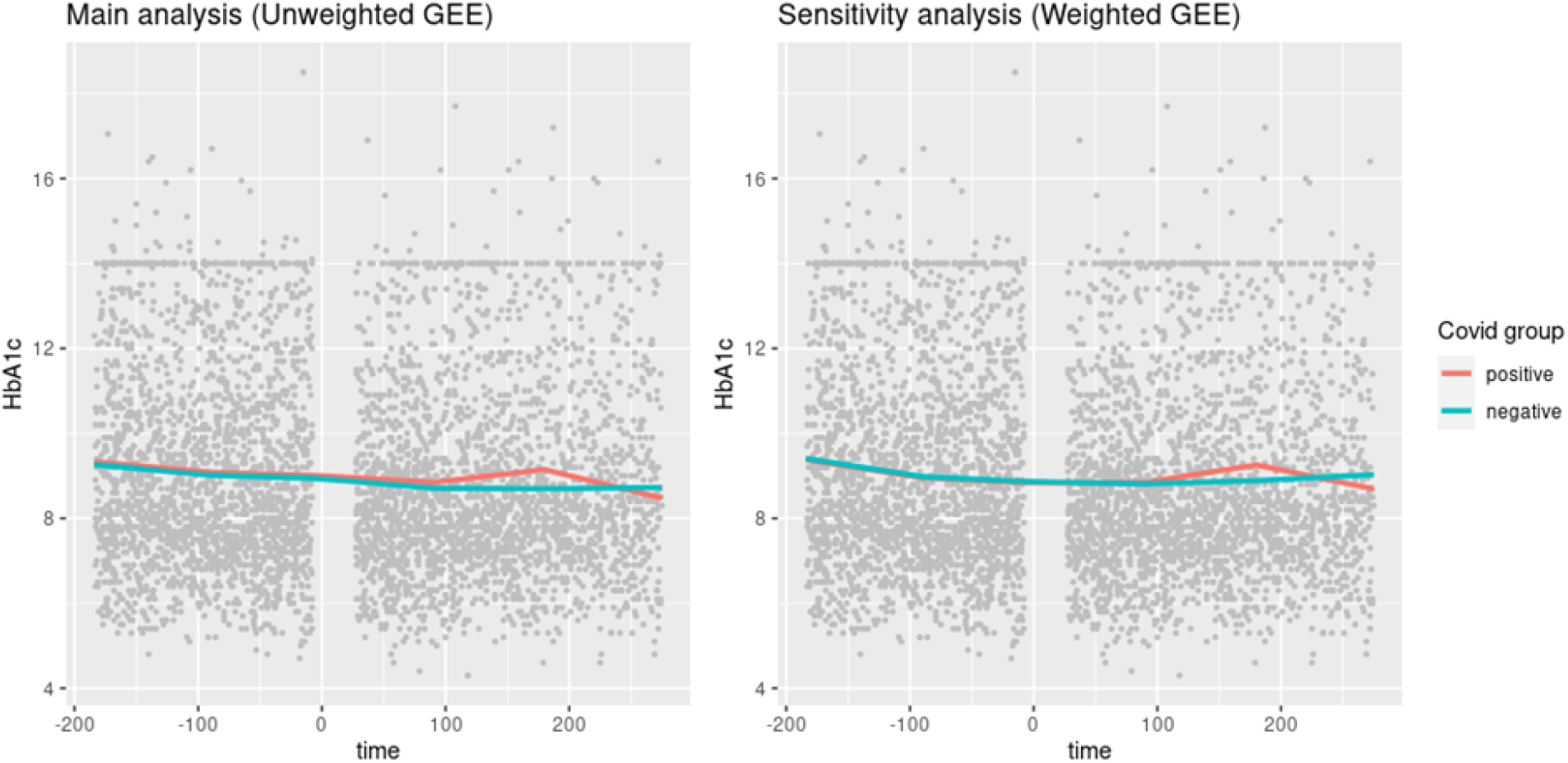
HbA1c trajectories before and after cohort entry in youth who tested positive or negative for COVID-19 from both main and sensitivity analysis.

## Discussion

This is one of the first studies in pediatrics examining the impact of COVID-19 infection on long term diabetes trajectory and outcomes for children with preexisting T1D. There was an increased risk of our composite outcome (worsening disease severity defined by increased utilization of the emergency department or hospitalization) in our Broad Cohort. While this did not reach statistical significance, this may be clinically significant for the population, since it DKA and SH both have serious health risks. The hazard ratio for the worsening composite outcome was higher prior to the Omicron wave, which mirrors findings from other studies demonstrating less severe disease and possibly lower risk of PASC following infection with omicron compared to alpha or delta variants(12; 39; 40).

Among those with HbA1c values 6 months to 7 days prior to cohort entry and a follow up A1c 28 days to 275 days after cohort entry, individuals positive for COVID-19 experienced a notable rise in HbA1c in the 3-to-6-month period after cohort entry. Also, the rate of decline of HbA1c > 6 months after cohort entry was steep when comparing our positive cohort to our negative cohort. This rise and fall could reflect sustained increases in glucose level after COVID-19 infection, and a return to baseline following recovery, which may also be due to dose adjustments at a clinical diabetes visit. Since HbA1c measurements reflect glucose control over the prior 3 months, acute illness following infection may also have contributed to these findings. However, the peak was noted in the later part of the 3- to 6-month window, consistent with the risk window for PASC, suggesting that glucose control remains challenging beyond the acute infection period. These findings extend our understanding of the longer-term impact of SARS-CoV-2 infection on glycemic control. Other studies have focused on the impact of infection during the acute illness, or in the immediate weeks after(41). However, we believe this is the first large-scale study to examine the direct impact of SARS-CoV-2 infection on glycemic control during the PASC risk window.

Our findings may have implications for understanding the long-term impact of COVID-19 in children with other chronic medical conditions. While most of the attention has been focused on “classic” symptoms of PASC that have been well described in adults, such as intermittent or prolonged respiratory, cardiac, or neurologic symptoms in individuals who were previously healthy, other potential manifestations of PASC are less well understood. Defining PASC in children is particularly challenging since PASC is often based on a lack of return to a usual state of health following acute COVID-19 illness. Children may be too young to articulate a change from baseline, and developmental baselines are ever-changing in children and youth. By widening the lens of PASC to include a broader range of manifestations, such as worsening disease trajectory following COVID-19 infection among children with chronic medical conditions, particularly given the variety of cells that can be infected by the SARS Co-V-2 virus(42), we may discover the burden of PASC in children is far greater than initially anticipated.

The strength of this work is the PEDsnet database which has multiple sites representing a diverse population around the country. PEDsnet derives its data from EHR rather than claims data which makes the data more complete. This is one of the largest studies examining PASC in children with type 1 diabetes with a well-matched comparison population. There are some limitations to this work. Since this data is based on EHR data, individuals diagnosed with COVID-19 outside of the participating centers may not be reflected. Similarly, health care utilization patterns for type 1 diabetes may have been altered by the pandemic, and there may have been residual differences between COVID-19 positive and negative cohorts despite balance on the observed covariates. We were also unable to accurately identify youth who had been vaccinated for COVID-19 prior to COVID-19 infection, which may be relevant if vaccination also provides protection against conditions associated with PASC. The decreased incidence of our composite outcome during the Omicron wave could indicate protection by vaccination or reflect a change in the severity of Omicron compared to original strains. Finally, given the relatively small number of youth with type 1 diabetes and COVID-19, our study may not be powered to detect significant differences.

## Conclusion

Children with type 1 diabetes diagnosed with COVID-19 were observed to have a transient increase in HbA1c in the 3 to 6 months following infection compared to COVID-19 negative cohorts, with substantial HbA1c declines after 6 months. As COVID-19 continues to circulate in the population, it is important to monitor the long-term disease trajectory in children with pre-existing T1D.

## Data Availability

All data produced in the present study are available upon reasonable request to the authors

## Disclosures

Dr Brill received support from Novartis and Regeneron Pharmaceuticals within in the last year

Dr. Chen receives consulting support from GSK.

Dr. Patel reports funding from the National Institute of Health and Bayer Pharmaceuticals

Dr. Rao reports prior grant support from GSK and Biofire and is a consultant for Sequiris.

Dr. Mejias reports funding from Janssen and Merck for research support; Janssen, Merck and Sanofi-Pasteur for Advisory Board participation, and Sanofi-Pasteru and AstraZeneca for CME lectures.

Dr. Jhaveri is a consultant for AstraZeneca, Seqirus, Dynavax, receives an editorial stipend from Elsevier and Pediatric Infectious Diseases Society and royalties from Up To Date/Wolters Kluwer.

Dr. Lee serves on the PASC Advisory Board for United Health Group.

All other authors have nothing to disclose.

## Acknowledgments

The authors gratefully acknowledge the contributions of Miranda Higginbotham, and we would like to thank the National Community Engagement Group(NCEG), all patient, caregiver and community Representatives, all patient, caregiver and community Representatives, and all the participants enrolled in the RECOVER Initiative.

## Notes

### Author Declarations

Childrens Hospital of Philadelphia Institutional Review Board designated this study as not human subjects research and waived informed consent.

